# Epidemic Model with Direct and Indirect Transmission modes and Two Delays

**DOI:** 10.1101/2022.03.16.22272508

**Authors:** F. Najm, R. Yafia, M. A. Aziz Alaoui, A. Aghriche

## Abstract

We propose an epidemic model in which all diseases are transmitted by direct transmission (infectives to susceptibles) and indirect transmission which occurs through shedding of virus by infectives and acquisition by susceptibles. The model takes into account the effect of latency period and time needed for a susceptible to become infective by indirect contact. Under the certain assumptions, the basic reproduction number of the model is identified from the direct and indirect basic reproduction numbers. The main goal is to analyse the asymptotic behaviours, global stability, bifurcation and to detecte the most sensitive parameters. Numerical simulations are carried out to illustrate the theoretical part.

## 1 Introduction and mathematical model

Infectious diseases are disorders caused by organisms such as bacteria, viruses, fungi or parasites and are serious public health problems for most countries in the world. Examples of such spreading diseases we cite influenza[16, 36], malaria [17], dengue [18], cholera [25], hantavirus[14], brucellosis [7], foot-and-mouth disease virus [32], infectious canine hepatitis virus [34], childhood diseases measles and rubella [8]. Some of these diseases are spread by organisms outside the host, and are highly important for the epidemiological dynamics and these organisms can be transmitted to the host by an indirect mode From this point of view a significant number of mathematical models suggested an indirect mode of transmission [5]. For diseases transmitted through indirect pathways with the pathogens to survive outside of its host has also been used to play a vital role in the transmission dynamics process of hantavirus [15, 28], cholera[5, 9, 23, 24, 29, 27, 33] and brucellosis [1, 12, 13, 21, 22]. In parallel to this mechanism, we find the direct transmission which postulates that the pathogen is acquired through an infectious contact with an infected individual. Some authors are developed some mathematical models to study the dynamics of infectious diseases by introducing a simple or multi-group mathematical models which involve both direct and indirect transmission. Mukandavire et al.[23, 24] introduced a dynamic model of cholera in Zimbabwe, Eisenberga et al. [9] proposed a cholera model in a patchy environment with water and human movement. In [10, 11, 20, 30, 31, 35] multi-group models have been introduced to describe the transmission dynamics of infectious diseases by using ordinary differential equations. In this paper, we propose an epidemic mathematical model which involve delays of direct and indirect transmissions. The model is given by the following delay differential equations system

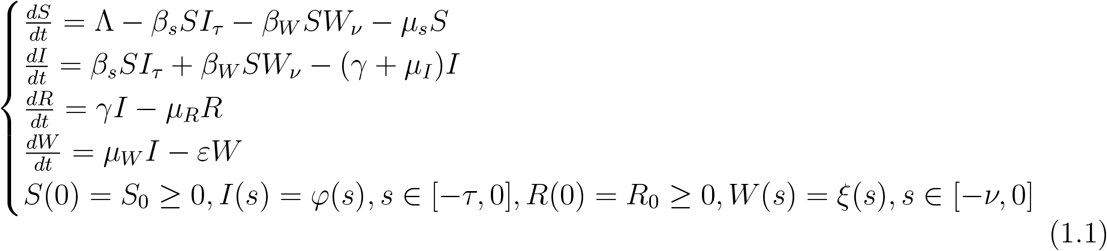

where *S, I* and *R* are the total number of susceptible, infected and recovered populations respectively. *W* is the concentration of virus caused by shedding. All parameters of the model are positive and are defined by, Λ is the birth rate parameter of S population, *β*_*s*_ is the transmission rate from I to S, *β*_*W*_ is the transmission rate from W to S, *μ*_*s*_ is the death rate of people S, *γ* is the recovery rate, *μ*_*I*_ is the death rate of I population, *μ*_*R*_ is the death rate of R population, *μ*_*W*_ is the shedding coefficients from I to 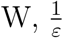 is the lifetime of the virus in W, *τ* is the latency period, *ν* is the time needed for a population *S* to become infected by indirect transmission

As the equation of recovered population *R* depends only on infected population *I*, it suffices to study the following reduced system:

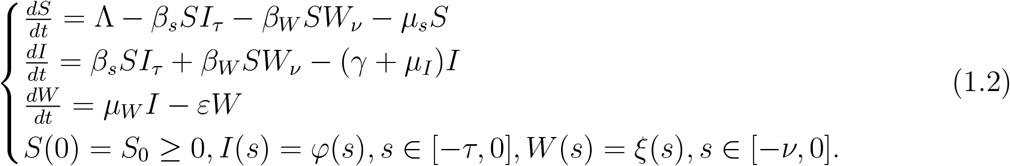

The rest of the paper is organized as follows. Section 2 deals with the positivity, boundedness of solutions and the existence of equilibria. The stability analysis without delays of the corresponding equilibria is established in Section 3 by applying Lyapunov method, we prove also the occurrence of a transcritical bifurcation for some critical value of the basic reproduction number *R*_0_. In Section 4 and 5, we consider the model with one and two delays respectively and we study the stability of equilibrium points. In Section 6 we prove how parameters can affect the basic reproduction number *R*_0_ by sensitivity analysis. Some numerical simulations are given in Section 7 in order to confirm and illustrate our analytical results. We end our paper by a conclusion.

## 2 Boundedness of solutions and equilibria

For system (1.2) to be biologically meaningful, it is necessary to show the non-negativity and boundedness of solutions.

### Proposition 2.1.

*The set* 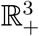 *is positively invariant with respect to system* (1.2) *with* (*τ* = *μ* = 0). *Furthermore, all solutions of* (1.2) *are uniformly bounded in the compact subset*.

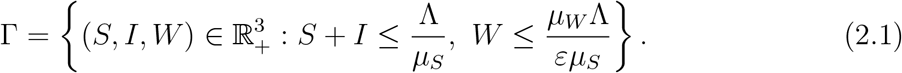

*Proof*. From the first equation of system (1.2), we have

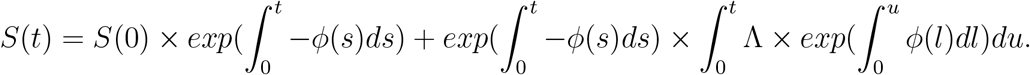

Thus *S*(*t*) *>* 0, ∀*t >* 0. To establish that ∀*t >* 0, *I*(*t*) *>* 0, *W* (*t*) *>* 0 whenever *I*(0) *>* 0, *W* (0) *>* 0, the above arguments can not be easily implemented. We then use an alternative trick. We Consider the following sub-equations related to the time evolution of variables

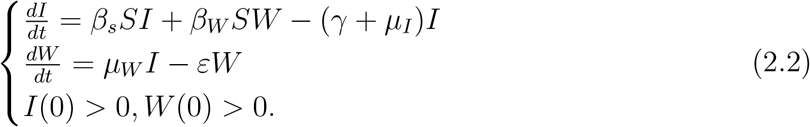

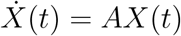

where 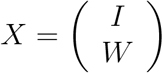 and 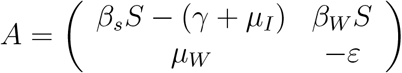

From the expression of *A*, its a Metzler matrix and its exponential is positive. Then we deduce the positivity of *I*(*t*) and *W* (*t*) whenever *I*(0) *>* 0 and *W* (0) *>* 0. This proves the positively invariant property of 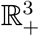 with respect to system (1.2).

Let *N* (*t*) = *S*(*t*) + *I*(*t*), then

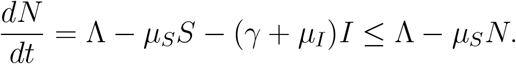

Hence,

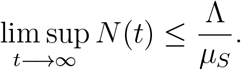

This implies that *S* and *I* are uniformly bounded in the region Γ. Furthermore, from the bound of *I* and the last equation of (1.2), it follows that

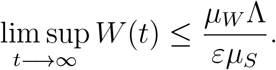

This guarantees the boundedness of *W*. This completes the proof. □

The equilibrium points of model (1.2) are obtained by solving the algebraic system obtained by cancelling all derivatives of *S*(*t*), *I*(*t*) and *W* (*t*). Thus:

1. The disease free equilibrium (DFE) is: 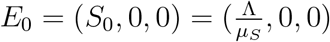.
2. The endemic equilibrium is: *E*_1_ = (*S*^*^, *I*^*^, *W*^*^), with its components given by

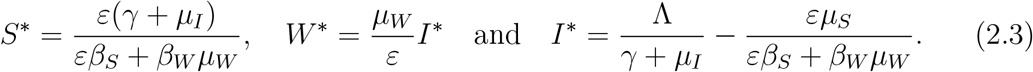

The endemic equilibrium *E*_1_ lies in the positive orthant if the basic reproduction number *R*_0_ of model (1.2) is greater than 1, it is given by:

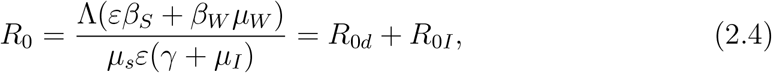

where 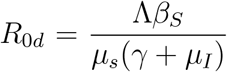 represents secondary infections caused directly by a single infective while 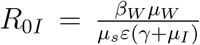 represents secondary infections caused indirectly through the environmental pathogen.

### Proposition 2.2.

- *If R*_0_ ≤ 1, *the system* (1.2) *has only the disease free equilibrium E*_0_ = (*S*_0_, 0, 0).
- *If R*_0_ *>* 1, *in addition to E*_0_ *the system* (1.2) *has a unique endemic equilibrium E*_1_ = (*S*^*^, *I*^*^, *W*^*^) *which is positive*.

## 3 Model without delay

Suppose *τ* = *ν* = 0, system (1.2) is written as

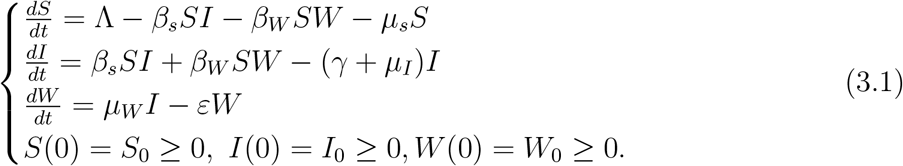

### 3.1 Stability Analysis

The aim of this section is to analyze the stability of the two equilibria *E*_0_ and *E*_1_.

Linearizing system (3.1) around an equilibrium point *E* = (*S, I, W*), we get the following jacobian matrix:

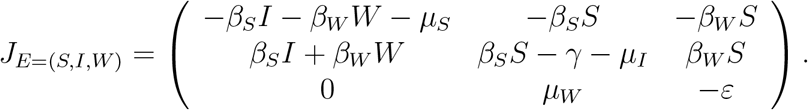

Replacing *E* by *E*_0_ and calculating the characteristic equation, we have

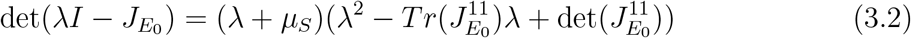

where

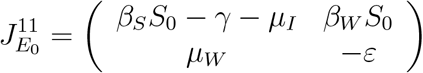

and

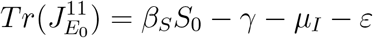

and

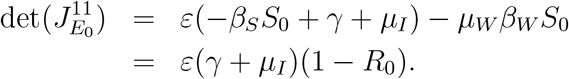

Then, we have the following result.

#### Proposition 3.1.

*If R*_0_ *<* 1, *the disease free equilibrium E*_0_ *is asymptotically stable and it becomes unstable if R*_0_ *>* 1.

*Proof*. If *R*_0_ *<* 1, then 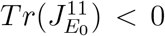 and 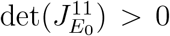, then the characteristic equation (3.2) does not admit a real strictly positive root.

If *R*_0_ *>* 1, then det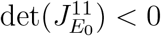. So, the characteristic equation (3.2) has at least one positive root.

#### Proposition 3.2.

*If R*_0_ *>* 1, *the endemic equilibrium E*_1_ *is asymptotically stable*.

*Proof*. Let *R*_0_ *>* 1 and we compute the characteristic equation associated to *E*_1_. The Jacobian matrix at *E*_1_ is given by

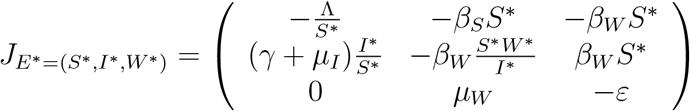

and the characteristic equation associated to *E*_1_ is as follows:

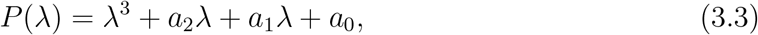

where

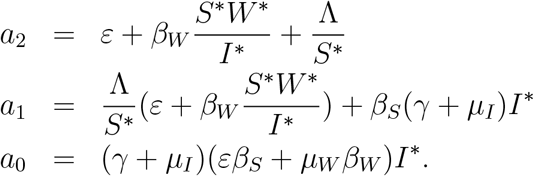

Note that if *R*_0_ *>* 1, then the coefficients *a*_0_, *a*_1_ and *a*_2_ are all strictly positive, then the polynomial *P* does not admit a real strictly positive root. □

#### Proposition 3.3.

*If R*_0_ *>* 1, *the endemic equilibrium E*_1_ *is globally asymptotically stable. Proof*. Define a Lyapunov functional as follows:

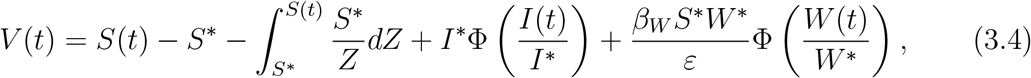

where Φ(*Z*) = *Z* − 1 − ln *Z >* 0, for *Z >* 0. It is obvious that Φ attains its strict global minimum at 1 and Φ(1) = 0. Then Φ(*Z*) *>* 0 and the functional *V* is nonnegative.

For convenience, let *ψ* = *ψ*(*t*) for any *ψ* ∈ {S, *I, W*}.

Differentiating *V* with respect to *t* along the solutions of (3.1), we obtain

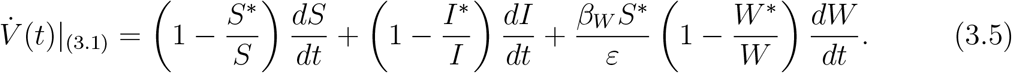

Using Λ = *μ*_*S*_*S*^*^ + (*γ* + *μ*_*I*_)*I*^*^ and *μ*_*W*_ *I*^*^ = *εW*^*^, we get

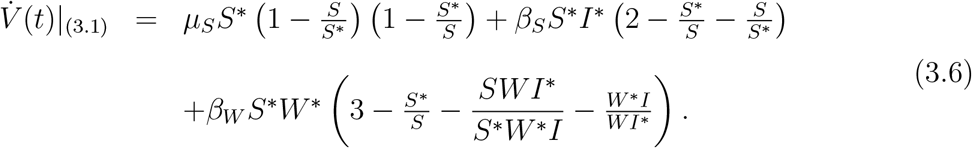

Thus,

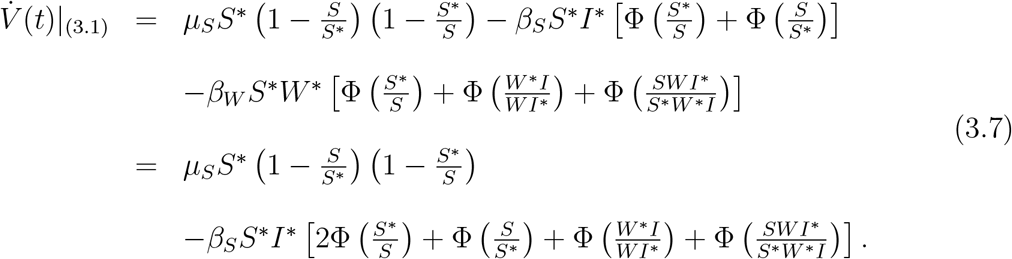

Since 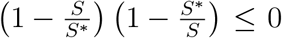 and Φ(*Z*) ≥ 0 for *Z >* 0, we deduce that 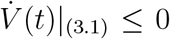 and the equality occurs at the endemic equilibrium *E*_1_. Consequently, the global asymptotic stability of *E*_1_ follows from LaSalle’s invariance principle [19]. □

### 3.2 Analysis at *R*_0_ = 1

In this section, we state and prove the following theorem which characterizes the occurrence of transcritical bifurcation at *R*_0_ = 1.

#### Theorem 3.4.

*The disease free equilibrium E*_0_ *changes its stability from stable to unstable at R*_0_ = 1 *and system* (3.1) *exhibits a transcritical bifurcation at bifurcation parameter value* 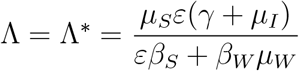.

*Proof*. Linearized matrix of system (3.1) around *E*_0_ at the bifurcation parameter value 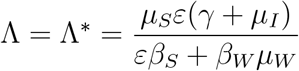 is given by

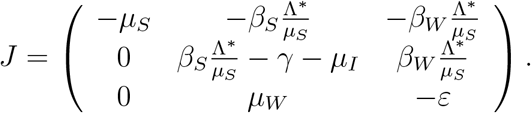

The matrix *J* has a simple zero eigenvalue at *R*_0_ = 1, (see characteristic equation 3.2) and the others eigenvalues have negative real part. At this stage the linearization techniques fail to conclude the behaviour of system (3.1). Center Manifold Theory is used to study the behaviour of non-hyperbolic equilibrium. Then from Theorem 1 of Castillo-Chavez and Song [3] the bifurcation constants *a*_1_ and *b*_1_ are given by

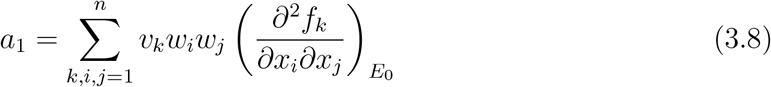

and

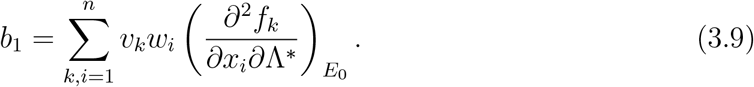

A right eigenvector associated with 0 eigenvalue is 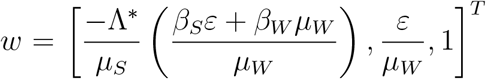 and the left eigenvector *v* satisfying *v*.*w* = 1 is 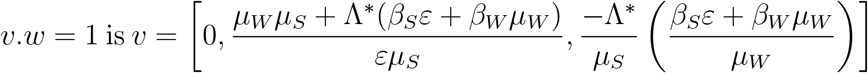.

Algebraic calculations show that 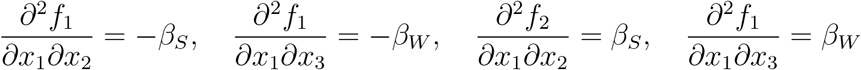 and 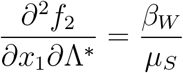.

The rest of the second derivatives appearing in the formula for *a*_1_ in (3.8) and *b*_1_ in (3.9) are all zero. Hence,

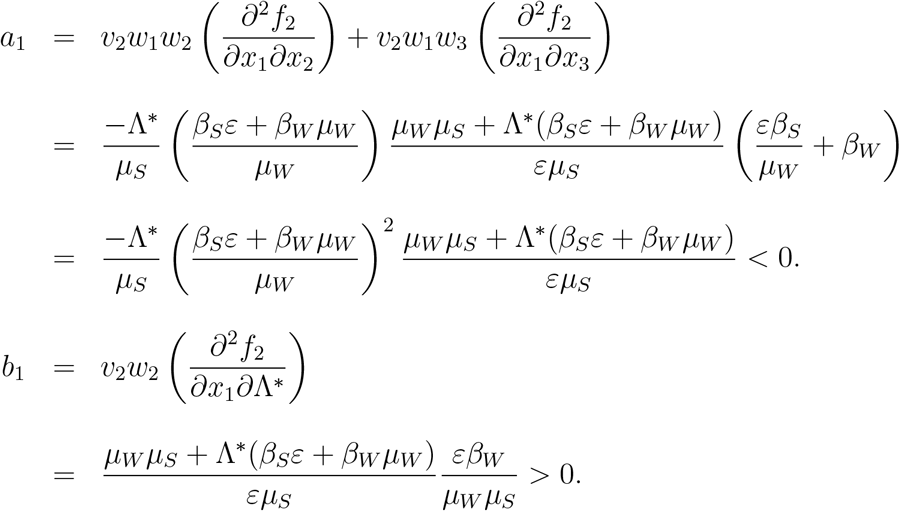

This shows that at *R*_0_ = 1, disease free equilibrium *E*_0_ changes stability from stable to unstable and endemic equilibrium *E*_1_ exists when *R*_0_ crosses the threshold value one. This emphasizes that, the system exhibts transcritical bifurcation at *R*_0_ = 1. □

## 4 Model with one delay *τ*

We suppose *τ >* 0 and *ν* = 0, system (1.1) is written as

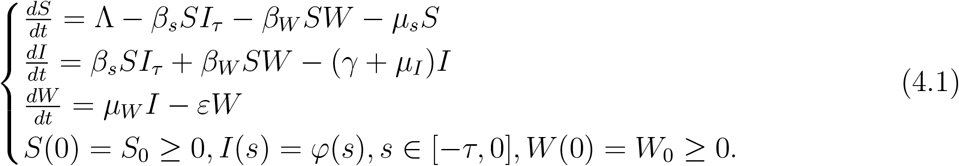

### 4.1 Local stability of disease free equilibrium

Linearizing system (4.1) around the disease free equilibrium *E*_0_ = (*S*_0_, 0, 0), we get

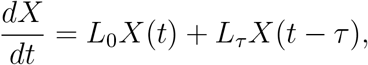

where *X* is a vector composed by state variables *S, I, W* and

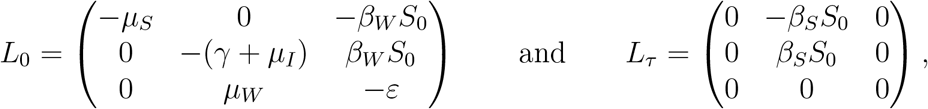

The associated characteristic equation is given by

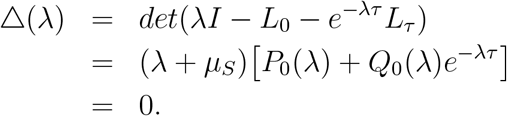

As *λ*_1_ = −*μ*_*S*_ is a root of the characteristic equation, so the study of stability of *E*_0_ is reduced to the study of the roots of equation

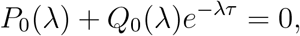

where

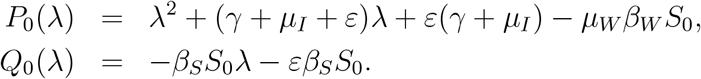

Define *F* by *F* (*y*) = |*P*_0_(*iy*)|^2^ − |*Q*_0_(*iy*)|^2^, (see [6]), then we have

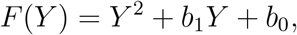

with *Y* = *y*^2^, and

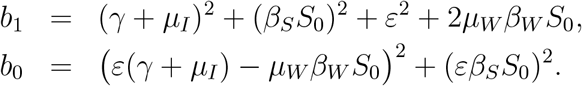

Since *b*_1_ *>* 0, *b*_2_ *>* 0, the function *F* does not admit a real strictly positive root. So there is no change in stability.

Therefore, we summarize the above discussions in the following result.

#### Proposition 4.1.

*The disease free equilibrium E*_0_ *is asymptotically stable for all τ >* 0.

### 4.2 Local stability of endemic equilibrium

In this section, we study the local asymptotic stability of the endemic equilibrium *E*_1_ = (*S*^*^, *I*^*^, *W*^*^) of system (4.1) considering latency period. The linearized system of (4.1) around *E*_1_ is given by:

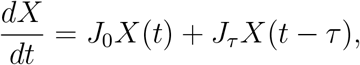

where

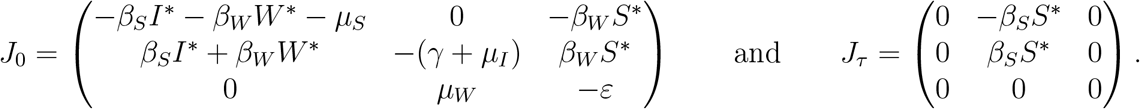

The associated characteristic equation to *E*_1_ is as follows

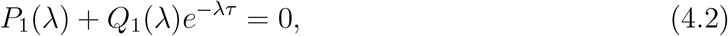

where

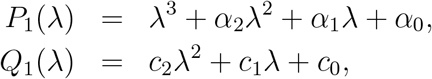

with

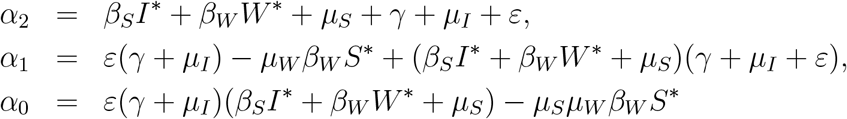

and

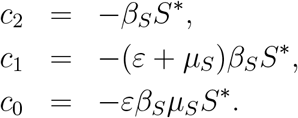

Since the endemic equilibrium *E*_1_ is asymptotically stable for *τ* = 0, (proposition (3.2)), by the continuity property, it’s still asymptotically stable for small *τ >* 0 [6, 2]. To obtain the switch of stability, one needs to find a purely imaginary root for some critical value of *τ*.

Let *iω* (*ω >* 0) be a root of Eq. (4.2), then we have

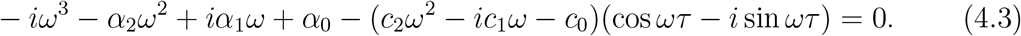

Separating the real and imaginary parts, we find

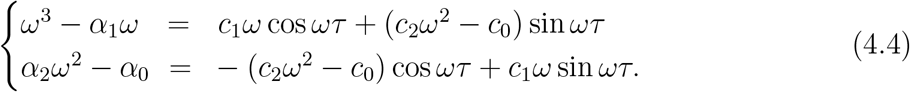

Adding up the squares of both the equations, we obtain

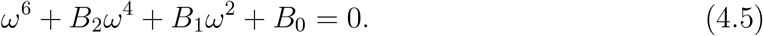

Let *z* = *ω*^2^, eq. (4.5) becomes

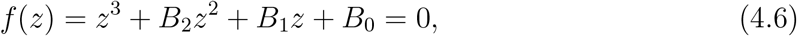

where

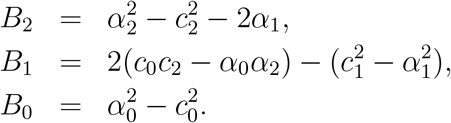

Let the hypothesis:

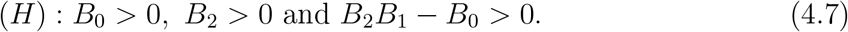

#### Proposition 4.2.

*If R*_0_ *>* 1 *and* (*H*) *is satisfied, then endemic equilibrium E*_1_ *is asymptotically stable for all τ >* 0.

*Proof*. The proof is deduced from the Routh-Hurwitz stability criterion [4]. □

## 5 Model with two delays *τ* = *ν >* 0

We consider *τ* = *ν >* 0, the model (1.2) is written as follows

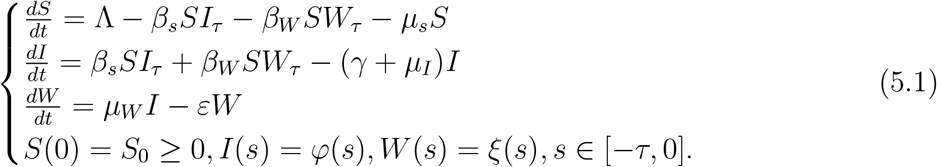

### 5.1 Local stability of *E*_0_

Linearising the system (5.1) around the disease free equilibrium *E*_0_, we get

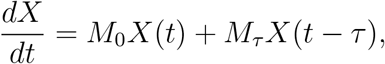

where,

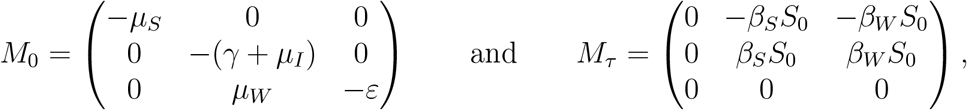

and the corresponding characteristic characteristic equation is as follows:

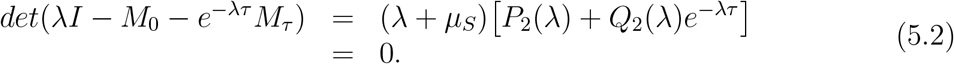

Since *λ*_1_ = −*μ*_*S*_ is a root of the characteristic equation, so the study of stability of *E*_0_ is reduced to the study of the roots of

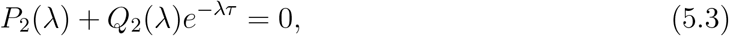

with

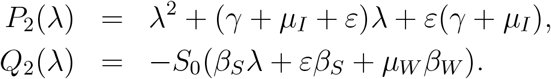

Define *G* by *G*(*y*) = |*P*_2_(*iy*)|^2^ − |*Q*_2_(*iy*)|^2^ and we have

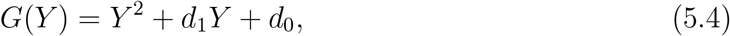

with *Y* = *y*^2^, and

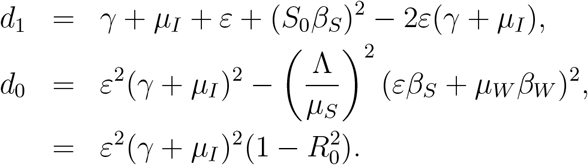

Note that, if *R*_0_ *>* 1 then *G*(0) = *d*_0_ *<* 0. As the function *G* is continuous and 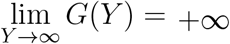. Then, equation (5.4) has at least one positive root, denoted by *Y*_0_. Consequently, the characteristic equation (5.2) has a pair of purely imaginary roots ±*iy*_0_. Hence the following result.

#### Proposition 5.1.

*The disease free equilibrium E*_0_ *is stable if R*_0_ ≤ 1 *and unstable when R*_0_ *>* 1 *for all time delay τ >* 0.

### 5.2 Local stability of *E*_1_

In this part, we study the local asymptotic stability of the endemic equilibrium *E*_1_ of system (5.1). By linearising system (5.1) at *E*_1_, we get

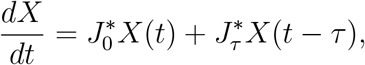

where

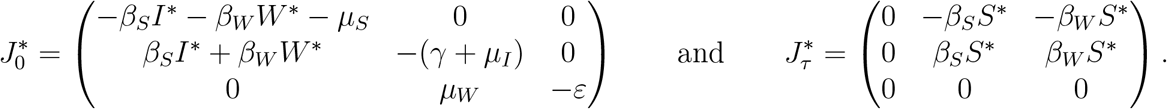

We get the following corresponding characteristic equation

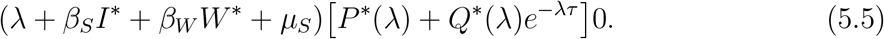

Since *λ* = −(*β*_*S*_*I*^*^ + *β*_*W*_ *W*^*^ + *μ*_*S*_) is a root of the characteristic equation, so the study of stability of *E*_1_ is reduced to the study of the roots of

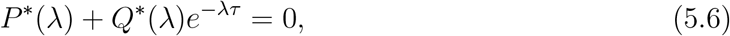

with

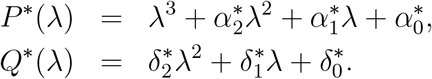

where

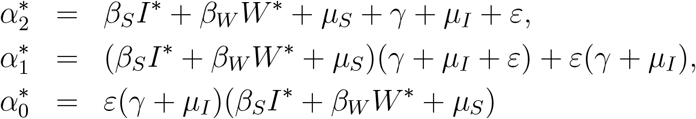

and

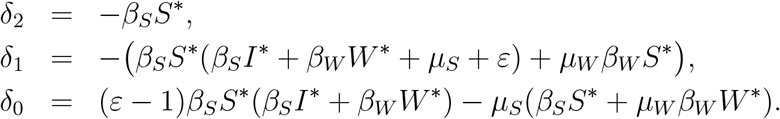

Since the endemic equilibrium *E*_1_ is asymptotically stable for *τ* = 0, by the continuity property, it’s still asymptotically stable for small *τ >* 0, (see proposition 3.2). To obtain the switch of stability, one needs to find a purely imaginary root for some critical value of *τ*.

Let *iω* (*ω >* 0) be a root of Eq. (5.6), then we have

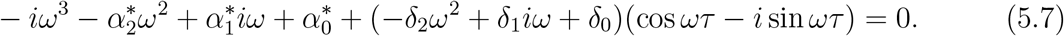

Separating the real and imaginary parts, we find

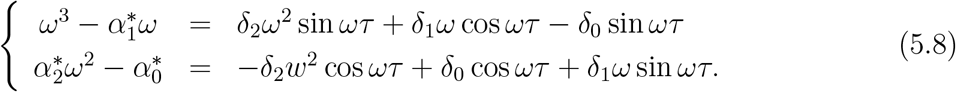

Adding up the squares of both the equations, we obtain

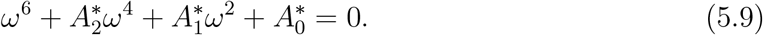

Let *z* = *ω*^2^, then equation (5.9) becomes

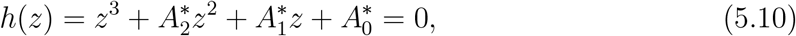

where

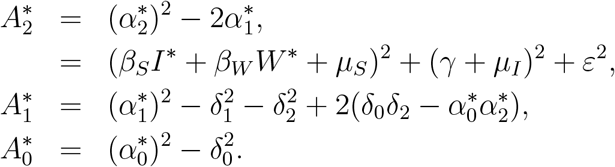

Since 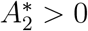, let the hypothesis:

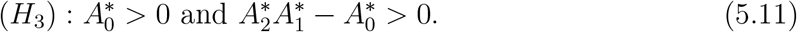

#### Proposition 5.2.

*If R*_0_ *>* 1 *and* (*H*_3_) *is satisfied, then endemic equilibrium E*_1_ *is asymptotically stable for all τ* = *ν >* 0.

*Proof*. The proof is deduced from the Routh-Hurwitz stability criterion [4]. □

## 6 Sensitivity Analysis

The sensitivity analysis for the endemic threshold (2.4) tells us how important each parameter is to disease transmission. This information is crucial not only for experimental design, but also to data assimilation and reduction of complex models [**?**]. Sensitivity analysis is commonly used to determine the robustness of model predictions to parameters values, since there are usually errors in collected data and presumed parameters values. It is used to discover parameters that have a high impact on the threshold *R*_0_ and should be targeted by intervention strategies. More accurately, sensitivity indices allows us to measure the relative changes in a variable when a parameter changes. For that purpose, we use the normalized forward sensitivity index of a variable with respect to a given parameter, which is defined as the ratio of the relative change in the variable to the relative change in the parameter. If such variable is differentiable with respect to the parameter, then the sensitivity index is defined as follows.

### Definition 6.1.

*[26] The normalized forward sensitivity index of R*_0_, *which is differentiable with respect to a given parameter θ, is defined by*

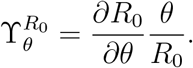

The values of the sensitivity indices for parameters values of Table 2, are presented in Table 1.

**Table 1:** Sensitivity of *R*_0_ evaluated for the parameters values given in Table 2

| Parameter | Sensitivity index | Index at parameters value |
| --- | --- | --- |
| $\Lambda$ | +1 | +1 |
| $\beta_S$ | $\frac{\varepsilon\beta_S}{\varepsilon\beta_S + \beta_W\mu_W}$ | 0.7500 |
| $\beta_W$ | $\frac{\mu_W\beta_W}{\varepsilon\beta_S + \beta_W\mu_W}$ | 0.25 |
| $\mu_S$ | -1 | -1 |
| $\gamma$ | $-\frac{\gamma}{\gamma + \mu_I}$ | -0.1818 |
| $\mu_I$ | $-\frac{\mu_I}{\gamma + \mu_I}$ | -0.8182 |
| $\mu_W$ | $\frac{\beta_W}{\varepsilon\beta_S + \beta_W\mu_W}$ | 8.3333 |
| $\varepsilon$ | $\frac{\varepsilon\beta_S}{\varepsilon\beta_S + \beta_W\mu_W} - \frac{\gamma}{\gamma + \mu_I}$ | 0.5682 |

Note that the sensitivity index may depend on several parameters of the system, but also can be constant, independent of any parameter. For example, $\Upsilon_{\theta}^{R_0} = +1$ means that

**Table 2:**
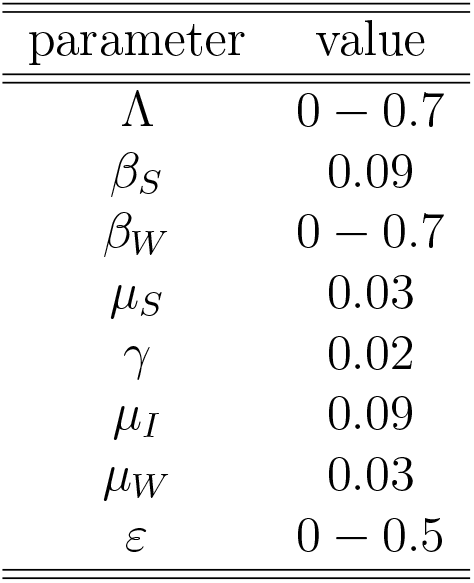
Parameters estimation

Note that the sensitivity index may depend on several parameters of the system, but also can be constant, independent of any parameter. For example, 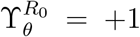 means that increasing (decreasing) *θ* by a given percentage increases (decreases) always *R*_0_ by that same percentage. The estimation of a sensitive parameter should be carefully done, since a small perturbation in such parameter leads to relevant quantitative changes. On the other hand, the estimation of a parameter with a rather small value for the sensitivity index does not require as much attention to estimate, because a small perturbation in that parameter leads to small changes.

From Table 1, we conclude that the most sensitive parameters to the basic reproduction number *R*_0_ of model (1.2) are Λ, *β*_*S*_, *μ*_*S*_, *μ*_*I*_ and *μ*_*W*_. In concrete, an increase of the value of *β*_*S*_ by (100%)will increase the basic reproduction number by 75% and this happens, in a similar way, for the parameters Λ and *μ*_*W*_. In contrast, an increase of the value of *μ*_*I*_ by (100%) will decrease *R*_0_ by 81.82% and this happens, in a similar way, for the parameter *μ*_*S*_.

## 7 Numerical simulations

In this section, we aim to provide a numerical simulation to substantiate the theoretical results established in the previous sections by using Matlab Software with the parameters given in table 2. Here, we consider *W* be a virtual reservoir population that fits to scaling of the population amounts and that is fitting by the infected (as stated) and decreases over time if no further input occurs.

## 8 Conclusions

In order to model the importance of the survival of pathogens outside the host, we proposed and investigated a mathematical model that describes the transmission dynamics of infectious diseases, which takes into account the effect of direct and indirect disease transmission and the period of latency. The well-posedness of the proposed model and the stability analysis of equilibria are rigorously studied. More precisely, we have established the existence, uniqueness, non negativity, and boundedness of solutions. By using appropriate Lyapunov functional and linearisation technique, we have proved that the first steady state *E*_0_ is asymptotically stable when *R*_0_ ≤ 1, which means that the disease dies out in the population. However, when *R*_0_ *>* 1, *E*_0_ becomes unstable and the model has an endemic steady state *E*_1_ which is asymptotically stable (Fig. 1). This leads to the persistence of disease. At *R*_0_ = 1 the transcritical bifurcation occurs. In the population when *R*_0_ *>* 1.

**Figure 1:**
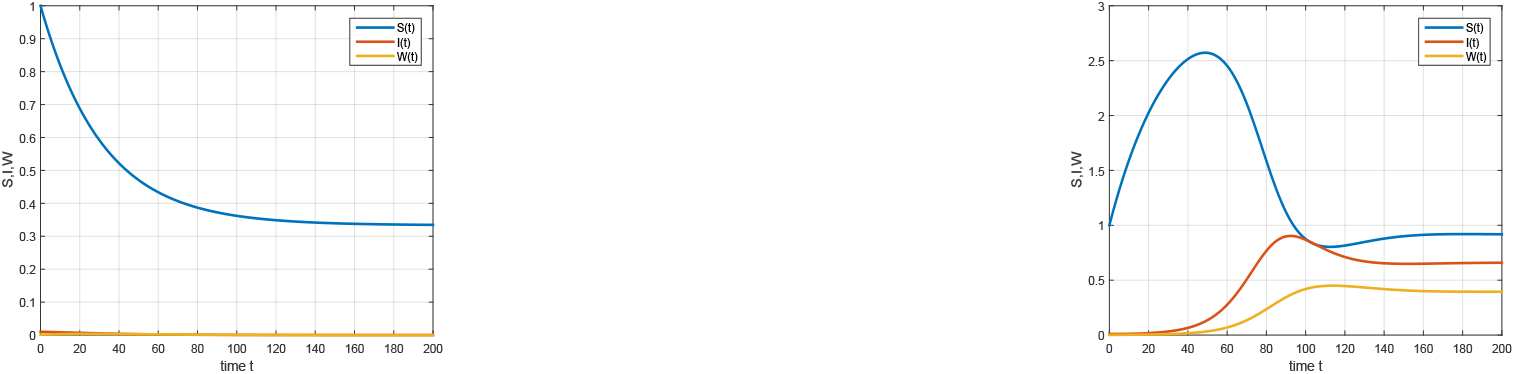
Stability of *E*_0_ and non existence of *E*_1_ for *τ* ≥ 0 and *ν* ≥ 0 (left) and Instability of *E*_0_ and stability of *E*_1_ for *τ* ≥ 0 and *ν* ≥ 0 (right).

The global stability is proved. From the sensitivity part and figures 2, 3, 4 and 5; the best strategy to stop the propagation of the infectious diseases is to decrease the concentration of environmental viruses.

**Figure 2:**
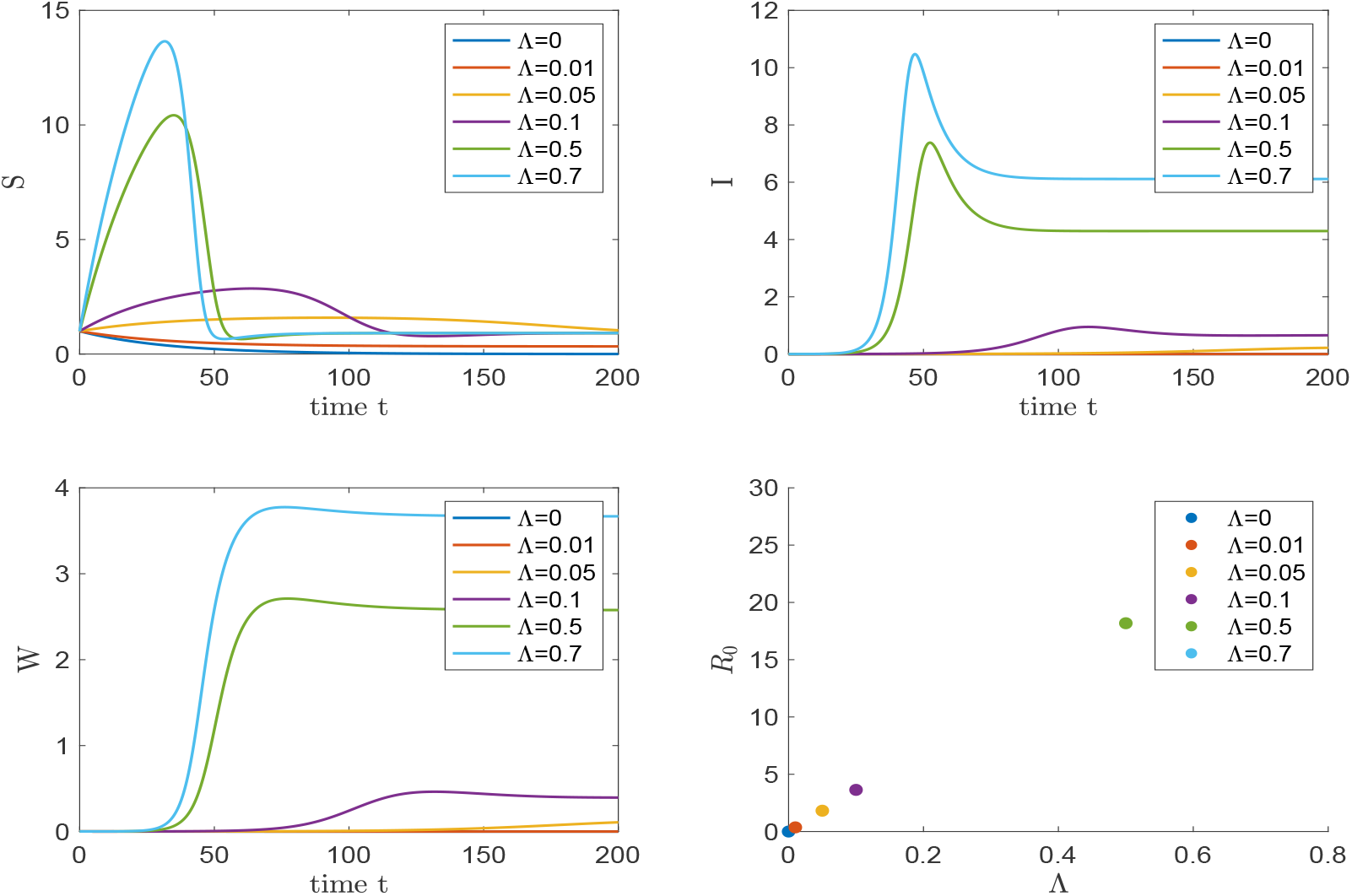
Temporal evolution of *S, I* and *W* for different values of Λ. If Λ increases, the number *R*_0_ increases which imply the increasing of the infected population and the virus concentration.

**Figure 3:**
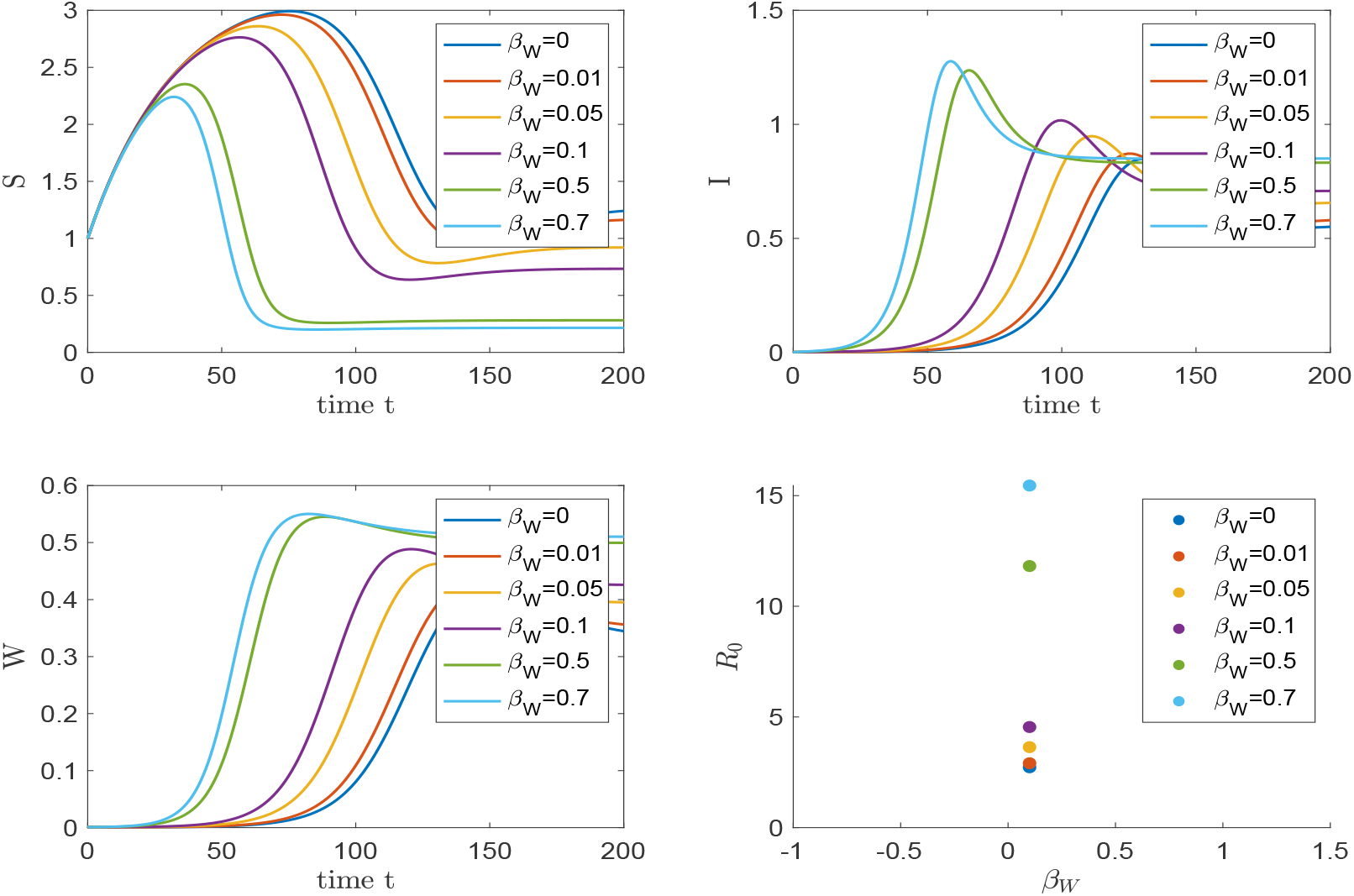
Temporal evolution of *S, I* and *W* for different values of *β*_*W*_. If *β*_*W*_ increases, the number *R*_0_ increases which imply the increasing of infected population and virus concentration.

**Figure 4:**
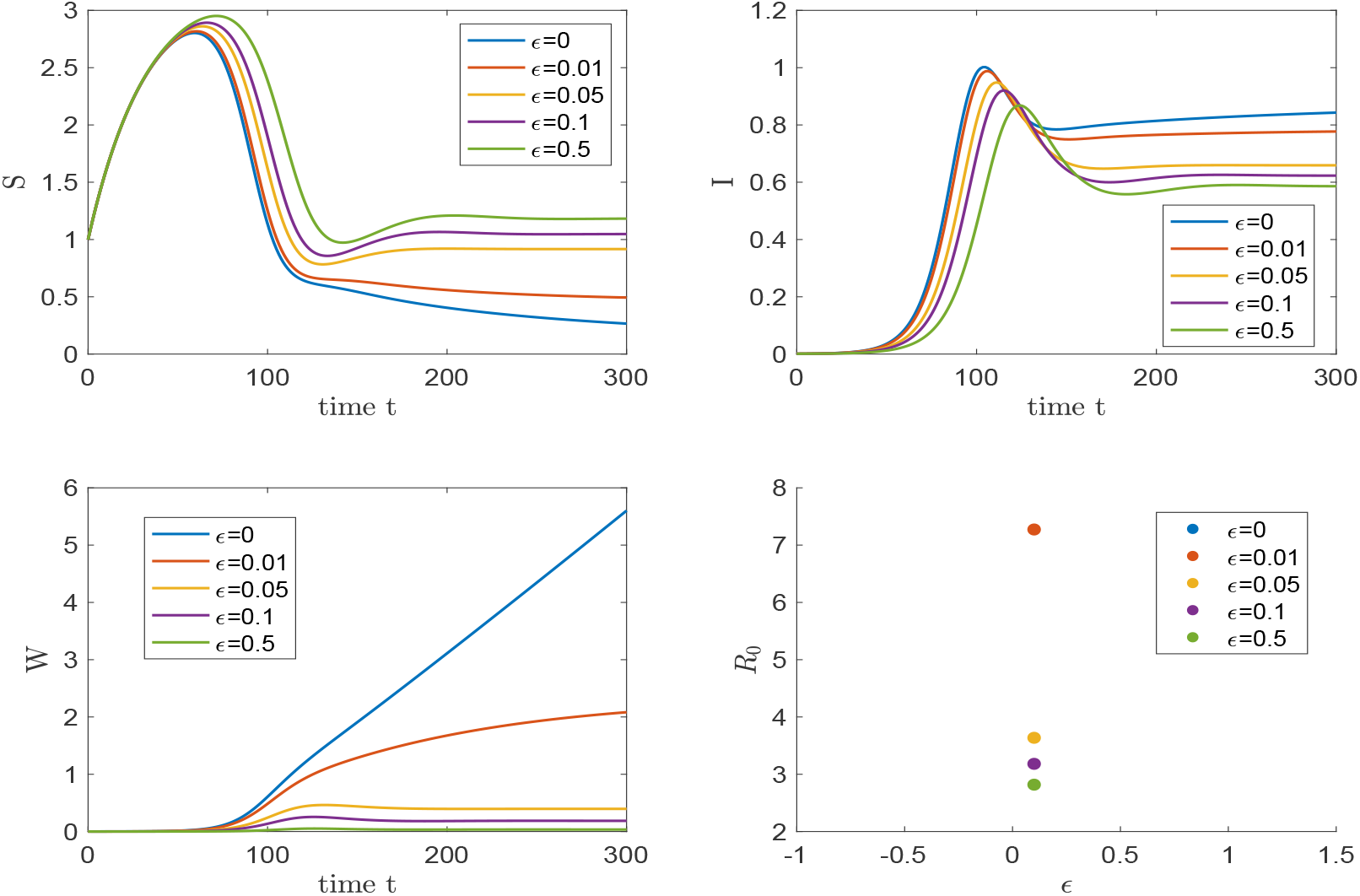
Temporal evolution of *S, I* and *W* for different values of *ϵ*. If *ϵ* increases, the number *R*_0_ decreases which imply the decreasing of infected population and virus concentration.

**Figure 5:**
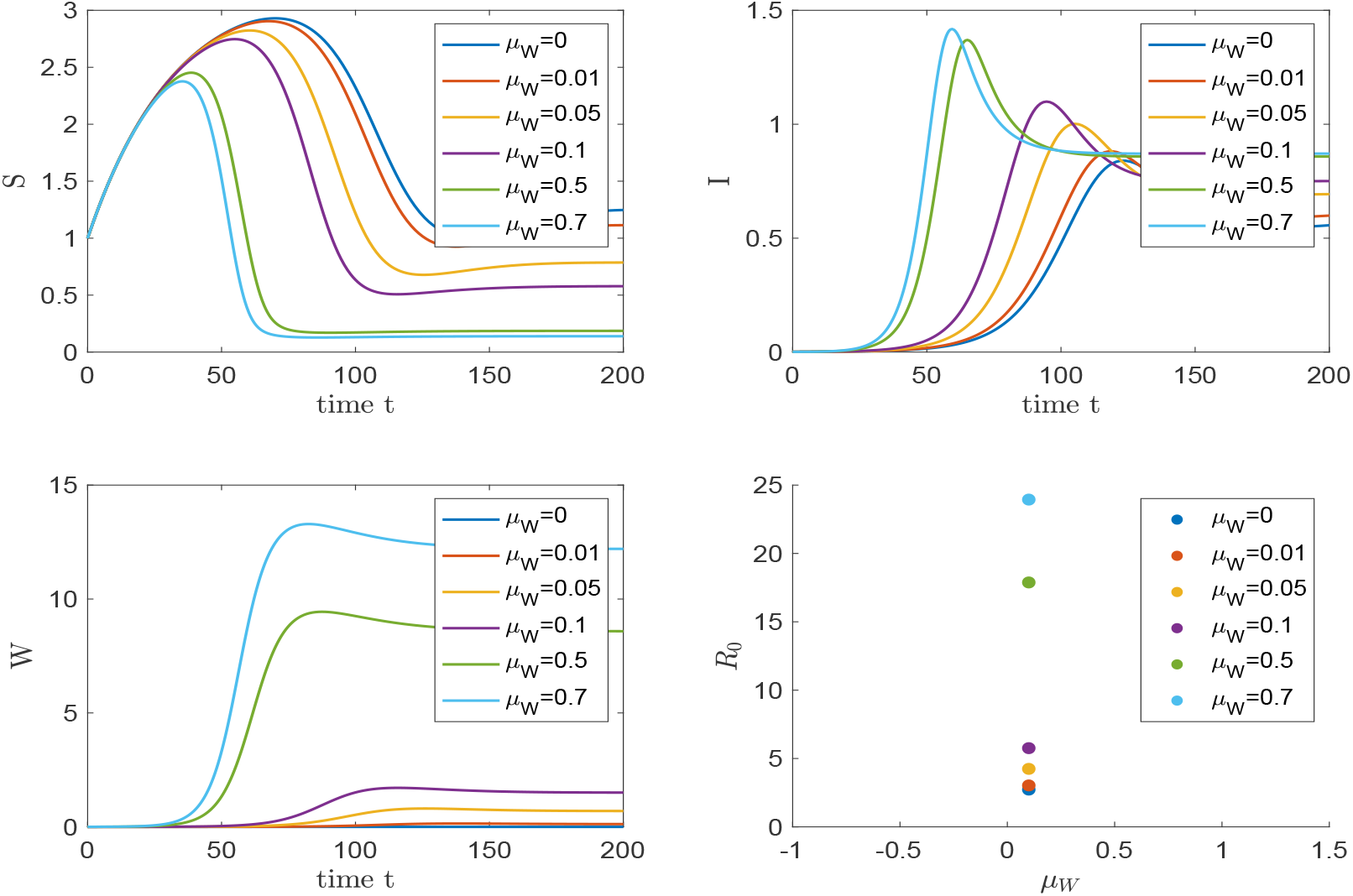
Temporal evolution of *S, I* and *W* for different values of *μ*_*W*_. If *μ*_*W*_ increases, the number *R*_0_ increases which imply the increasing of infected population and virus concentration.

## Data Availability

No data

